# Social inequalities in human mobility during the Spanish lockdown and post-lockdown in the Covid-19 pandemic of 2020

**DOI:** 10.1101/2020.10.26.20219709

**Authors:** Alberto Hernando, David Mateo, Ignacio barrios, Angelo Plastino

## Abstract

Many countries established strong population lockdowns as a response to the pandemic of COVID-19 in 2020. While these measures proved efficient in stopping the spreading of the virus, they also introduced collateral effects in the economies of these countries. We report in this work that the imprints in mobility of both the lockdown and post-lockdown on the Spanish population are measurable by means of the daily radius of gyration using mobile phone data. We cross mobility with economic data segmented by average salary per person so as to find large inequalities between low- and high-income populations. Indeed, low-income populations typically show a 17% higher radius of gyration than high-income ones during pre-lockdown (8.1 km vs. 6.9 km). However, this relative difference grows to a maximum during lock-down (3.3 km vs. 900 m) since most of the essential workers (carriers, nurses, supermarket cashiers, farmworkers, etc.) belong to the first segment. Post-lockdown shows reversed inequality in the weeks during summer vacations as high-income populations multiplied their pre-lockdown radius by 70% as a rebound effect driven by leisure, while low-income populations recovered their normal pre-lockdown radius. This period is correlated with an extraordinary increase in the number of new Covid cases, which stabilized after the holyday weeks once at the so-called *new normal*. We find that this new normal emphasizes the pre-lockdown inequalities in mobility between low- and high-income population, increasing the inequality up to a 47%. These results show the relevance of devising measures that could account for potential collateral inequalities.

## 1. Introduction

The World Health Organization (WHO) declared the outgoing COVID-19 international crisis as a pandemic on March 11, 2020 [1]. To that date, over 118,000 positive cases had been reported in over 110 countries and territories around the world. The spread risk became real as the number of cumulated confirmed cases grew to 31.7 million in October over all continents in all continents, with more than 1.1 million deceases [2].

The first positive case of COVID-19 in the Spanish territory was detected on January 31, 2020 [3]. It was followed by many others in the following weeks. As of March 9, the number of cases exploded in the Madrid forcing authorities to take partial mobility restrictions. However, positive cases had already been detected in all Spanish regions and the government declared a lockdown with what is called a State of Alarm on 15th March [4]. Many other countries adopted similar measures, with strong restrictions in mobility that proved to be efficient to control the virus-spreading [5-8]. However, they ultimately led to multiple economic and social collateral effects [9].

A recent study showed that the centralized structure of the country’s communication network contributed to export positive cases from Madrid to all other Spanish provinces [10]. Indeed, the number of visits per capita from travellers between Madrid and every province in the weeks before the lockdown strongly correlates with the peak of sickness incidence, mortality, and antibody prevalence in the first pandemic wave. Three months of lockdown with a strict reduction in both intracity and intercity mobility helped to reduce the new number of cases to a minimum. This was the expected goal of these measures as the central role of human mobility in epidemics has been extensively considered in the past [11-18] and more recently for COVID-19 in particular [10,19,20].

It is also known that mobility patterns also correlates with the socio-economic structure of cities. Indeed, urban space segregation modulates the social opportunities of its inhabitants and may generate inequalities of access to resources and jobs [21-23]. While these inequalities have been reported in the past, it is still a question how the pandemic will affect the social equilibrium the post-pandemic era, the so-called new normal.

The explicit correlation between mobility and the spreading of COVID-19 in Spain was obtained after analysing Call Detail Records (CDR) data from a major mobile operator. CDR data has been widely used to understand mobility in general [24,25] and recent studies are showing its convenience for tracking the correlations with the pandemic effects in particular [26,27]. The radius of gyration is an efficient metric used to quantify the mobility of a region by place of residence which has been show also useful to measure the effects of the lockdown in the pandemic [20,26,28].

In view of the potential of the above cited tools to understand the effects of the pandemic, we present in this work measurements of the radius of gyration crossed with the average salary per person for all Spain in 2020 to find potential socioeconomic inequalities during the pandemic. To this aim, we use CDR data from 15 million users homogeneously distributed in the Spanish territory, which represent one of the largest datasets ever reported. We find that the year 2020 can be divided into four definite periods according to collective mobility patterns: i) pre-lockdown, showing the pre-existing reference patterns in mobility; ii) lockdown, characterized by a drastic drop in mobility; iii) summer holidays, displaying a rebound effect motivated by leisure trips; and iv) new normal, with a general reduction in mobility as compared with the pre-existing pattern. We then correlate the radius of gyration with the average salary per person, confirming the above-mentioned pre-existing inequalities in the pre-lockdown era and finding new ones during the lockdown and post-lockdown stages. In particular, we find that the relative inequality between populations of low- and high-salaries exhibits a maximum during the lockdown. This may be attributed to the fact that many of the essential workers belong to the low-salaries sector. These newer inequalities were not fully understandable in the new normal context, as pre-existing inequalities grew due to telecommuting. In addition, we find a strong parallelism between (1) the four periods defined by mobility and (2) the evolution of the number of new COVID-19 cases. Such parallelism reaffirms the relevance of mobility so as to understand the evolution of the pandemic.

In brief, our results will demonstrate that any preventive measure against the pandemic should account for potential collateral inequalities to avoid extra instabilities in the years to come after the pandemic

### 2. Data and Methods

We use place of residence as proxy to correlate mobility with average salary. The Spanish National Institute of Statistics (INE) provides official geo-referenced statistical indicators of the level and distribution of household incomes at the municipal and inframunicipal levels. This dataset is constructed from INE’s geographical and social information together with tax data from both the Spanish Tax Agency and Regional Treasuries. We use in this work the Average Income per Person at the inframunicipal level of censal section, openly obtained from these sources (both data and geographical zonification) at Ref. [29].

Mobility is measured using Call Detail Records from one of the major telecommunication operators in Spain. The data set includes 13 million users in a time-span of 42 weeks, from 6^th^ of January 2020 to 18th of October 2020. The user’s residence is defined as the most common censal section visited from 10pm to 6am and it is updated monthly. The sample is scaled per each censal section by a scaling factor as to reproduce the total population amount provided by INE. The daily radius of gyration per user RG is measured as [25]

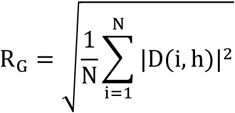

where N is the number of events per each user and D(i, h)is the geodesic distance between the i-th daily event and the user’s homeplace. We assign a weekly value per user as the weekly average (7 days from Monday to Sunday). We obtain the long-tailed distribution depicted in Fig. 1 when aggregating for all users. The mode of the distribution is located at the lowest bin of 1 km with a median of 3.2 km and a mean value of 8.0 km. The big difference between median and mean is indeed typical of these long-tailed distributions.

**Fig. 1.**
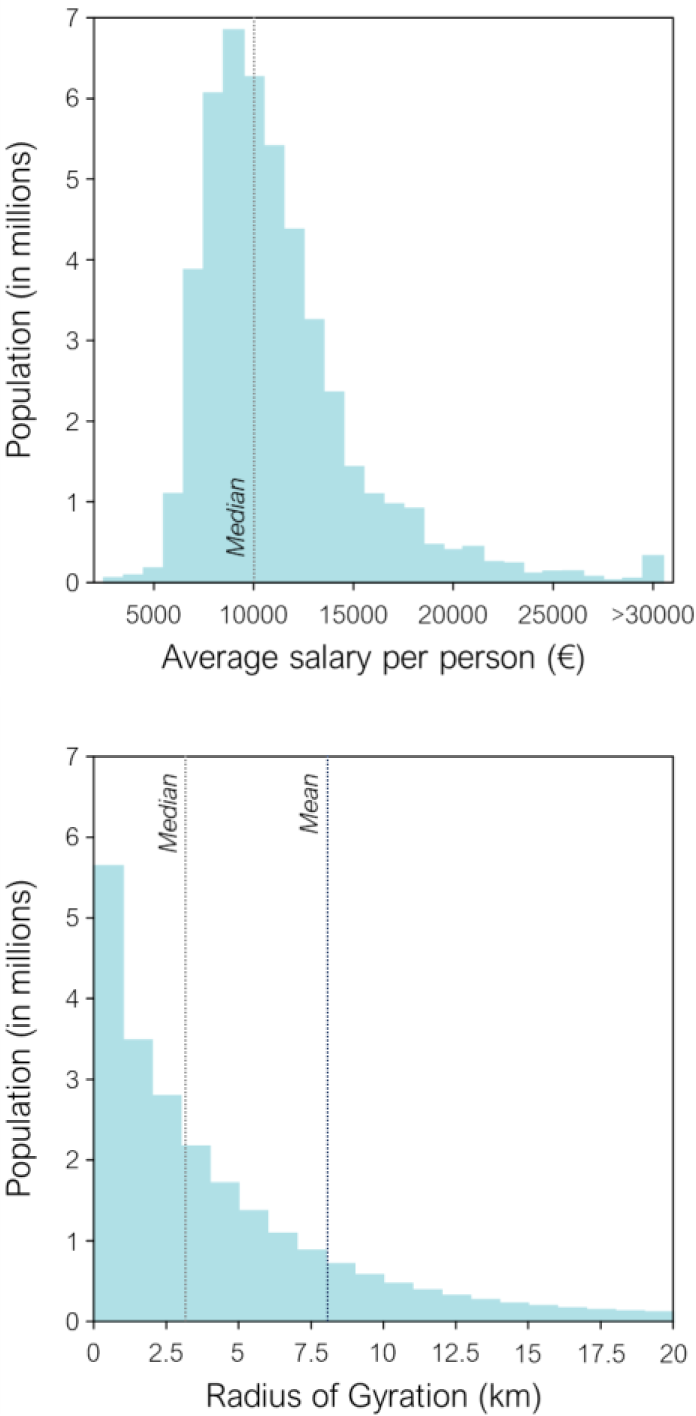
Distribution of the average salary per person assigned by place of residence and distribution of the radius of gyration for all Spanish population.

We assign to each user the average salary per person corresponding to the censal section assigned as residence and aggregate the values into regular intervals of 1000€, rounding to the nearest thousand. The last bin for highest salaries aggregates all values greater or equal to 30,000€, as provided by INE. We obtain the distribution of average salary showed in Fig. 1. The distribution displays a positive skew, typical of salary distributions.

We also compare the evolution of mobility with the evolution of new COVID-19 cases per week. We use the updated dataset provided by the World Health Organization [2].

## 3. Results

To correlate average salaries with the mean radius of gyration, we consider all the users belonging to a given bin of salary and thus obtain the mean value of the radius per week. We find the evolution displayed in Fig. 2a from weeks 1 to 42 of 2020. Remarkably enough, we can clearly define four periods according to collective mobility patterns:

**Fig 2.**
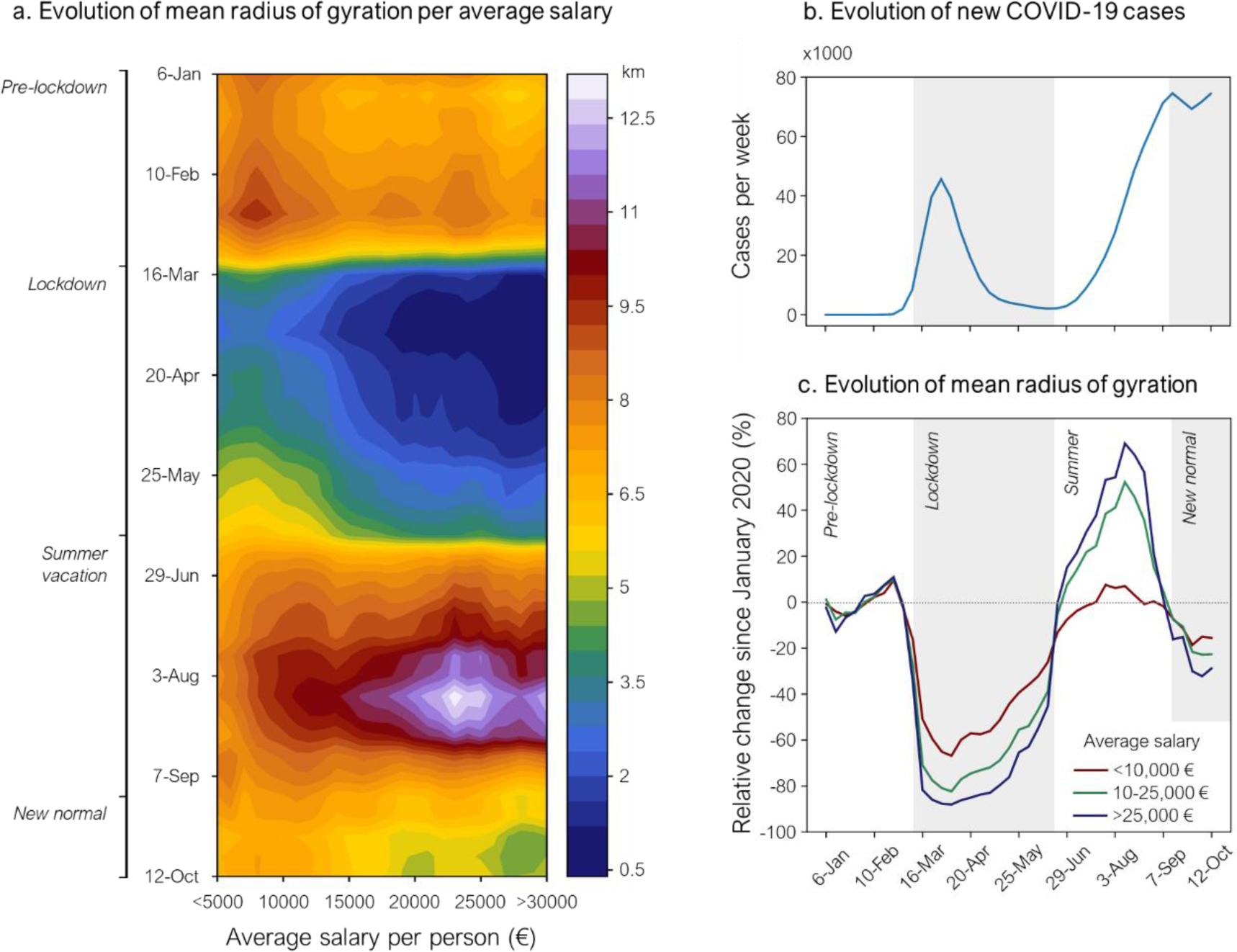
**a**. Evolution of mean radius of gyration per average salary for every week of 2020 until week 42 (starting at Monday 12th October). We can differentiate four clear periods according to mobility patterns: pre-lockdown, lockdown, summer holydays and new normal (see text for details). **b**. Evolution of the number of new COVID-19 cases for the same period. Remarkably, the same four periods are fitted by the curve. **c**. Evolution of the mean radius of gyration per average salary aggregated. It refers to i) 50% of the population with lower salary (<10,000€), ii) the top highest salaries (>25,000€) and, iii) the rest (10-25,000€).

### I. Pre-lockdown: from 1^st^ of January to 15th of March

This period represents the former normal before the pandemic. It is characterized by a higher mean radius for low-income populations (8.1 km) that for high-income populations (6.9 km) as shown in Fig. 3 in detail. This difference represents an inequality, since low-income populations spent 17% more on their daily trips than high-income populations, mostly in commuting via both private or public transport.

**Fig. 3.**
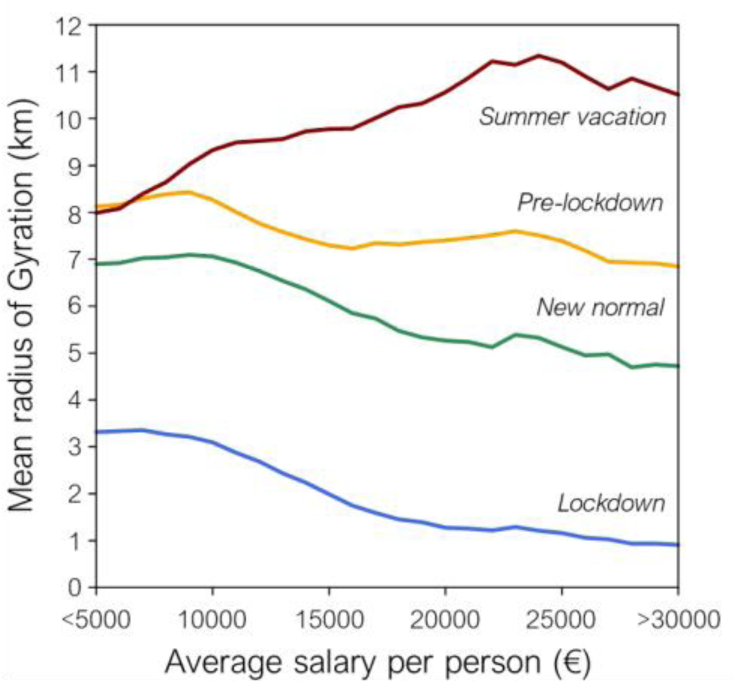
Mean radius of gyration for the four periods defined by collective mobility patterns (see text for details).

### II. Lockdown: from 15th of March to 20th of June

Spanish lockdown was one of the most restrictive ones in Europe, motivated by the fast growth of cases in the previous weeks. Its imprint in collective mobility is clear: the radius of gyration dropped down to 900 m for the highest-income population. For the lowest, it dropped down to 3.3 km only (Fig. 3). This represents the highest inequality in relative terms between income segments (266%). The underlying reason is that essential workers continued their work during the lockdown, and most of these workers belong to the lowest income range (carriers, nurses, supermarket cashiers, farmworkers, etc). Such fact also indicates an extra exposure to COVID-19 in this period. Fortunately, the lockdown worked as expected and the number of COVID-19 cases declined to a virtual zero (Fig. 2b).

### III. Summer holydays: from 20th of June to 14th of September

Shortly after the end of the lockdown the summer holydays affected millions of people. This period is characterized by a rebound effect for that half of the population belonging to the higher salary segment, whose radius grew up to a 70% with respect the value at pre-lockdown. Despite the pandemic, national tourism expanded in fast fashion with a peak in August, as reported by specialized local journals [30]. However, the half of the population belonging to the lowest salary range did not experience this rebound, as they recovered the value of their radius at the pre-lockdown stage. Most of these low-income workers continued with their daily commuting to work or did not travel during their holydays. In parallel, the number of COVID19 cases grew again every week, reaching a maximum at the end of this period, in a second wave that arrived weeks earlier than in the rest of Western Europe [who].

### IV. New normal: from 14^th^ of September to 18^th^ of October

The new normal is characterized, among other factors, by telecommuting (TC). We can see the TC-effect in a general decrease in the radius of gyration. However, this is more easily visible for higher-income populations, for which the mean radius fell down to 4.7 km, while the pertinent lower-income population value is 6.9 km. This fact represents an inequality of 47%, much higher than that at the pre-lockdown era. The number of COVID-19 cases stabilized in this period at large values. The number of cases continued growing after this period and new measures and restrictions in people’s mobility were ordered by the Spanish authorities.

## 4. Discussion and conclusion

The radius of gyration is an efficient metric to compare the average mobility of the population between different locations. A pre-existent inequality existed before the pandemic between low- and high-income populations, as measured by their radius of gyrations. The increase of telecommuting is a useful measure to prevent exposure to COVID-19, but it is not egalitarian since not any type of work can ascribe to this modality. Indeed, the post-lockdown radius of gyration tell us that mobility decreases more in the upper-half range of salaries than in the lower-half range. This difference escalated from 17% to 47%, meaning that the population with the lowest salaries expend in average a 47% more in their daily displacements (in cost and time) that the population with the highest salaries. Many of these low-income workers are considered essential, meaning that they were more exposed to the virus during the lockdown. In addition, this segment of the population was commuting as usual during the holiday period, not exhibiting any statistical difference with the patterns observed at the pre-lockdown era.

The cumulation of such inequalities may introduce an undesired variable to the complex situation of the pandemic. Future strategies for new lockdowns or preventive measures in the new normal should account for these collateral inequalities and prevent or compensate for potential stress in the population. It is important to measure the effects of these strategies with updated data since collective behaviour can change from one week to another in the new normal, as seen in our results.

## Data Availability

The information on mobility are available upon request with the permission of Kido Dynamics SA. The data on average salary per person and geographical borders for cesal sections can be downloaded from Ref. [29]. The data on number of COVID cases can be downloaded from Ref. [2].

## Data availability statement

The information on mobility are available upon request with the permission of Kido Dynamics SA. The data on average salary per person and geographical borders for cesal sections can be downloaded from [29]. The data on number of COVID cases can be downloaded from [2].

## Acknowledgements

The authors thank Jose Javier Ramasco from IFISC (UIB-CSIC) and Marta Gonzalez from University of Berkeley for useful discussions.

